# Genotype-first approach to the detection of hereditary breast and ovarian cancer risk, and effects of risk disclosure to biobank participants

**DOI:** 10.1101/2020.06.29.20139691

**Authors:** Liis Leitsalu, Marili Palover, Timo Tõnis Sikka, Anu Reigo, Mart Kals, Kalle Pärn, Tiit Nikopensius, Tõnu Esko, Andres Metspalu, Peeter Padrik, Neeme Tõnisson

**Affiliations:** Estonian Genome Center, Institute of Genomics, University of Tartu, Tartu, Estonia; Institute of Molecular and Cell Biology, University of Tartu, Tartu, Estonia; Institute for Molecular Medicine Finland, University of Helsinki, Helsinki, Finland; Dept. of Clinical Genetics, United Laboratories, Tartu University Hospital, Tartu, Estonia; Hematology and Oncology Clinic, Tartu University Hospital, Tartu, Estonia; Institute of Clinical Medicine, University of Tartu, Tartu, Estonia

**Keywords:** population-based biobank, genotype-first approach, return of results, BRCA1/2, hereditary breast and ovarian cancer, population screening

## Abstract

Genotype-first approach allows to systematically identify carriers of pathogenic variants in *BRCA1*/*2* genes conferring a high risk of familial breast and ovarian cancer. Participants of the Estonian biobank have expressed support for the disclosure of clinically significant findings. With an Estonian biobank cohort, we applied a genotype-first approach, contacted carriers and offered return of results with genetic counseling. We evaluated participants’ responses to and the clinical utility of the reporting of actionable genetic findings. Twenty-two of 40 contacted carriers of 17 pathogenic *BRCA1*/*2* variants responded and chose to receive results. Eight of these 22 participants qualified for high-risk assessment based on National Comprehensive Cancer Network criteria. Twenty of 21 counseled participants appreciated being contacted. Relatives of 10 participants underwent cascade screening. Five of 16 eligible female *BRCA1*/*2* variant carriers chose to undergo risk-reducing surgery, and 10 adhered to surveillance recommendations over the 30-month follow-up period. We recommend the return of results to population-based biobank participants; this approach could be viewed as a model for population-wide genetic testing. The genotype-first approach permits the identification of individuals at high risk who would not be identified by application of an approach based on personal and family histories only.

## INTRODUCTION

The rapidly increasing volume of genomic data at the population scale imposes an intensified need for best practices to guide and standardize the way how clinically significant, but potentially unexpected, genetic findings from population-based research cohorts are handled. The human genomics community needs to address questions such as how to anticipate and manage such genetic events, and how to communicate such unexpected findings.

Several biobanks in Australia, Northern Europe, and the United States have applied a genotype-first approach, in which individuals carrying clinically significant variants are re-contacted and offered returns of results.^1–4^ Being irrespective of family history and the presence of medical indication, this approach varies greatly from common practice in clinical settings, where considerations regarding genetic analysis are based on personal or familial histories. Evidence for participants’ reactions to unexpected results is limited, but suggests that people tend to appreciate being given actionable information about their health risks.^1,3,5–8^

Although the general consensus supports the return of clinically significant findings in clinical and research settings, numerous challenges must still be faced, especially in the research context.^3,6,8^ One such challenge involves the provision of genomic risk information to apparently healthy individuals in a manner that improves disease risk prediction, prevention, diagnosis, and therapy. Procedural guidelines are currently limited, largely due to a dearth of conclusive studies on the clinical and psychological impacts of such disclosure.

The unique legislative setting in Estonia permits the re-contacting of Estonian Biobank (EstBB) participants for specific research and prospective studies. The Estonian Human Genes Research Act^9^ and the broad consent given by all biobank participants allow us to conduct valuable research on the impact of the communication of potentially unexpected genomic findings to research participants.

Given its high prevalence in the EstBB dataset, carrier screening for hereditary breast and ovarian cancer (HBOC; MIM #604370, #612555) was selected for this study. In the general population, about 5–10% of all breast cancer and 10–15% of all ovarian cancer cases can be attributed to variants in the *BRCA1* (MIM #113705) and *BRCA2* (MIM #600185) genes, which can explain about half of breast/ovarian cancer aggregation in families.^10–12^ The prevalence of clinically significant genetic variants in *BRCA1*/*2* is about 1/400 in the general population, but can vary significantly depending on the characteristics of specific study cohorts.^13^ Women with an inherited *BRCA1* pathogenic variant have a 65– 72% lifetime risk of breast cancer development, and a 37–62% risk of ovarian cancer development; *BRCA2* pathogenic variant carriers are expected to have lifetime risks of 45– 85% for breast cancer and 11–23% for ovarian cancer by the age of 80 years.^14–16^

In Estonia, guidelines for the identification of high-risk individuals are based largely on those of the National Comprehensive Cancer Network (NCCN); *BRCA1/2*-associated HBOC is suspected in individuals with personal or family histories.^17^ The main approach of identifying risk variants in multiplex families with high frequencies of breast and ovarian cancer is common in clinical contexts, but has been shown to miss a large percentage of high-risk individuals.^18^ In this study, we applied a genotype-first method with data from the population-based EstBB to systematically identify individuals at high risk of HBOC, regardless of phenotypic heterogeneity, incomplete penetrance, or lack of family history.

## MATERIALS AND METHODS

### Study cohort

The EstBB is a population-based biobank managed by the Institute of Genomics at the University of Tartu.^19^ It currently contains data from more than 200,000 participants, representing almost 20% of Estonia’s adult population. All participants have provided broad written consent, which encompasses the provision of samples for future research use and the collection of electronic health records from national registries and databases. EstBB participants can opt out of future re-contact regarding participation in additional research projects or the receipt of research results.

Data for this study are from a sub-cohort of 17,679 EstBB participants recruited between 2002 and 2011. At the time of analysis, sequencing data were available from a genome sequencing (GS) and an exome sequencing (ES) dataset for 4,594 individuals. Array-based genotype data (Illumina Infinium HumanCoreExome and OmniExpress-12 beadchip microarrays) were available for the remaining 13,085 individuals. Sequenced reads were aligned to the GRCh37 human reference genome assembly. After filtering and exclusion of duplicates between the two datasets, the final GS and ES sets contained 2,240 and 2,354 unique samples, respectively. Sequenced variants were annotated with the Variant Effect Predictor^20^ (version 87; Gencode v19 on GRCh37.p13) and ANNOVAR.^21^ GS and ES data preparation is described in detail in the Supplementary Materials and Methods.

For long-range phasing, we used high-coverage (30×) GS and array-based genomic data from 2,240 and 13,085 EstBB participants, respectively. The GS panel was phased using the read aware phasing model of SHAPEIT v2.r837,^22^ with the *read-quality* and *base-quality* parameters set to 20. Genotype data were phased with SHAPEIT v2.r837 using the default parameters. The GS and array-based data were merged, leaving only overlapping single nucleotide variants for further analysis. We calculated pairs of individuals based on shared identity by descent using PLINK v1.9,^23^ with the *min* parameter set to 0.4. Haplotypes of calculated pairs were then visualized using R software,^24^ and the graphs generated were assessed visually. All cases of putative alternative allele carriage were analyzed further by Sanger sequencing.

### Evaluation of variant pathogenicity

We extracted all *BRCA1*/*2* coding variants from the GS/ES data, and cross-referenced those with likely pathogenic (LP) or known pathogenic (KP) classification and minor allele frequencies < 0.5% with the ClinVar,^25^ VariantValidator^26^ and dbSNP 153 databases.^27^ For estimation of the pathogenicity of all LP/KP variants, several *in silico* prediction algorithms were used. Further details of variant detection and evaluation of variant pathogenicity are provided in the Supplementary Materials and Methods.

We compiled a list of carriers of 17 LP/KP *BRCA1*/*2* variants^28^ and validated each by Sanger sequencing. For confirmation, independent blood samples collected at participants’ first visits were tested by an ISO-certified (ISO 15189:2012, ISO 9001:2015) diagnostic laboratory (Asper BioGene Ltd, Tartu, Estonia).

### Return of results procedure

The procedural framework for the communication of unexpected genetic findings with high clinical relevance to EstBB participants was developed with participants’ rights and interests at the forefront. A previously developed procedure for the return of results and provision of clinical support^5^ was implemented with a few modifications (Figure 1). This framework is consistent with the precepts of Estonian Human Genes Research Act,^9^ and the protocol for this study was approved by the Research Ethics Committee of the University of Tartu.

**Figure 1.**
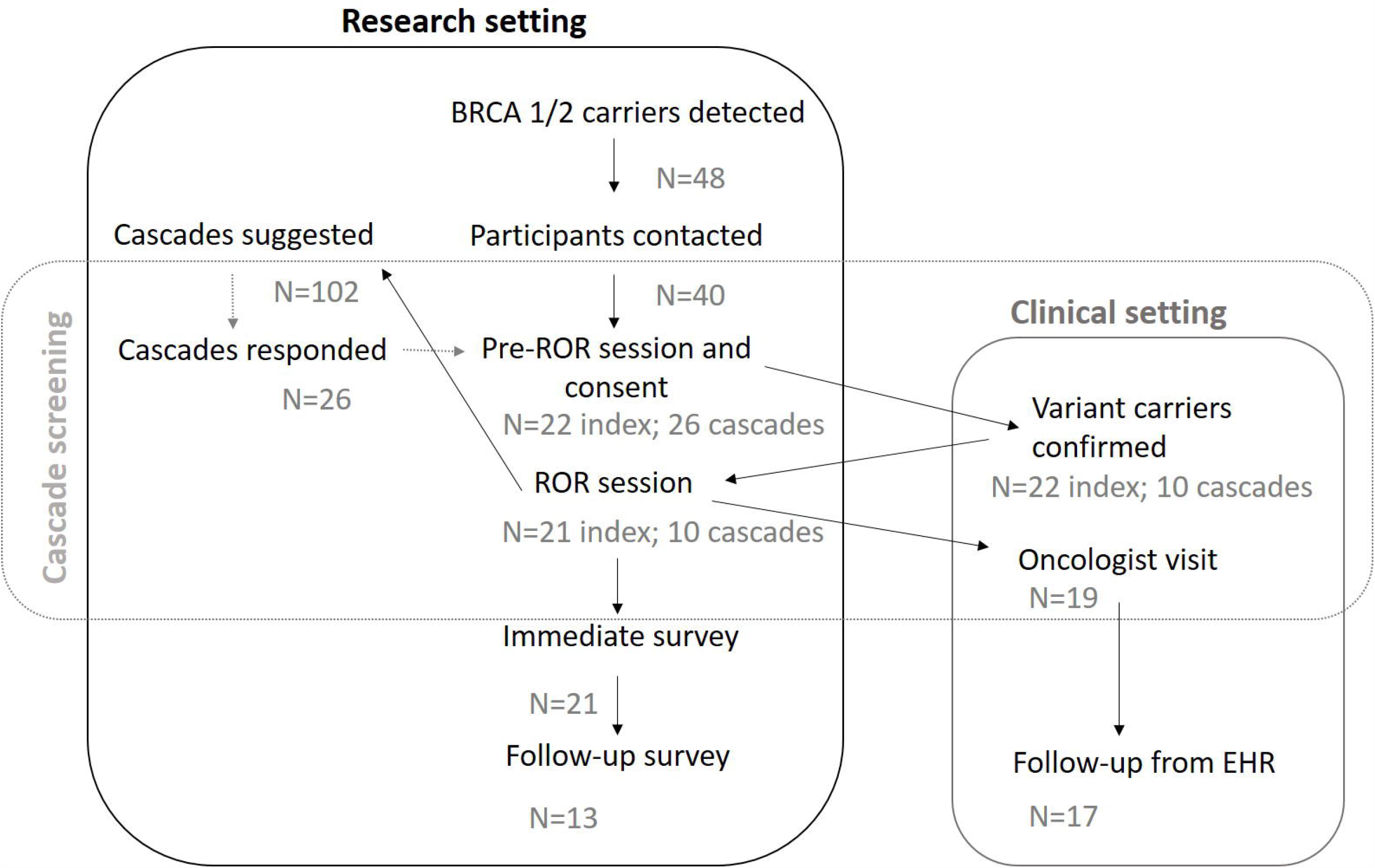
Framework for the return of results. Most steps were conducted in the research setting. Female LP/KP variant carriers were referred to a collaborating oncologist.

The framework includes procedures for the contacting of variant carriers without immediate disclosure of their genetic status: the sending of an initial contact letter (Supplementary Material S1.1), acquisition of project-specific informed consent, independent validation of the finding in a certified diagnostic laboratory, disclosure of genetic risk accompanied by genetic counseling, and collaboration with an oncologist for the development of a personalized participant surveillance plan. Cascade screening was included to identify additional high-risk individuals in carrier families. Information letters were provided to index individuals to hand out to the respective family members invited to cascade screen (Supplementary Material S1.2).

### Follow-up

Data on participants’ responses to the receipt of results about clinically significant findings were gathered using immediate and long-term feedback surveys, developed based on findings from analogous previous studies.^5,29–31^ The first survey, administered immediately after the disclosure of genetic results, included questions about participants’ satisfaction, understanding, and psychological responses.^30^ The second survey, mailed to participants 6 months later, included questions about decision regret,^31^ perceived personal control and coping,^32^ psychological adjustment,^30^ communication, support, and reported health behavior and healthcare utilization. Cascade screening participants were asked to fill in a survey immediately after the disclosure of clinically significant findings.

To facilitate the provision of effective clinical support for high-risk variant carriers identified in a research setting, all female participants carrying LP/KP *BRCA1*/*2* variants were referred directly to a collaborating clinical oncologist. These participants’ ongoing medical management was reviewed via electronic health records from the collaborating hospital and national imaging and e-health databases. Depending on the dates of participants’ first visits, follow-up periods ranged from 12 to 30 months (January 2017– July 2019).

## RESULTS

### Identified *BRCA1*/*2* genetic variants

Forty-eight individuals in the study cohort were identified as carriers of 17 LP/KP *BRCA1*/*2* variants (Table 1). Eighteen carriers were identified from the GS cohort, 19 were identified from the ES cohort, and additional 11 carriers were identified through long-range phasing of genotyping array data. Ten of the 17 variants were in *BRCA1* (in 35 participants) and seven were in *BRCA2* (in 13 participants). Fifteen (88%) of the variants had been classified in ClinVar previously: 14 KP variants had 3* status and one KP variant had 2* status. One LP variant had pending status in the Breast Cancer Information Core database (not actively curated).^33^ Based on GS and ES data, the prevalence of high-risk *BRCA1*/*2* variants was 0.80% (1/124), first degree relatives excluded. For extended details refer to Supplementary Materials and Methods.

**Table 1.** Identified HBOC-associated genetic variants.

| Gene | LP/KP<br>variant | rsID | cDNA position<sup>1</sup> | Protein position | GS/ES<sup>2</sup> | LRP<sup>3</sup> | AC_EST<sup>4</sup> |
| --- | --- | --- | --- | --- | --- | --- | --- |
| BRCA1 | KP | rs80356898 | c.1687C>T | p.Gln563* | 1 | 0 | 1 |
| BRCA1 | KP | rs28897672 | c.181T>G | p.Cys61Gly | 1 | 0 | 1 |
| BRCA1 | KP | rs80357282 | c.1840A>T | p.Lys614* | 1 | 0 | 1 |
| BRCA1 | LP | novel | c.2178delT | p.Pro727Glnfs*9 | 1 | 0 | 1 |
| BRCA1 | KP | rs80357711 | c.4035delA | p.Glu1346Lysfs*20 | 11 | 5 | 16 |
| BRCA1 | KP | rs80357508 | c.4065_4068delTCAA | p.Asn1355Lysfs*10 | 1 | 0 | 1 |
| BRCA1 | KP | rs80357305 | c.4258C>T | p.Gln1420* | 1 | 0 | 1 |
| BRCA1 | KP | rs80356860 | c.5117G>A | p.Gly1706Glu | 1 | 0 | 1 |
| BRCA1 | KP | rs80357906 | c.5266dupC | p.Gln1756Profs*74 | 7 | 4 | 11 |
| BRCA1 | LP | rs483353102 | c.5534_5539delinsCCAGTGCC<br>AGGACAGCAGG | p.Tyr1845Serfs*39 | 1 | 0 | 1 |
| BRCA2 | KP | rs80358622 | c.37G>T | p.Glu13* | 1 | 0 | 1 |
| BRCA2 | KP | rs397515636 | c.3975_3978dupTGCT | p.Ala1327Cysfs*4 | 1 | 0 | 1 |
| BRCA2 | KP | rs886040543 | c.469_470insT | p.Lys157Ilefs*26 | 1 | 0 | 1 |
| BRCA2 | KP | rs80359584 | c.6405_6409delCTTAA | p.Asn2135Lysfs*3 | 1 | 0 | 1 |
| BRCA2 | KP | rs80359112 | c.8572C>T | p.Gln2858* | 4 | 2 | 6 |
| BRCA2 | KP | rs1555288494 | c.9097_9098insT | p.Thr3033Ilefs*11 | 2 | 0 | 2 |
| BRCA2 | KP | rs876661242 | c.9381G>A | p.Trp3127* | 1 | 0 | 1 |

HBOC, hereditary breast and ovarian cancer; LP, likely pathogenic; KP, known pathogenic; GS, genome sequencing; ES, exome sequencing; LRP, long-range phasing.
<sup>1</sup>Provided for *BRCA1* ENST00000357654 (NM\_007294.3) and *BRCA2* ENST00000380152 (NM\_000059.3).
<sup>2</sup>*n* = 4,594 individuals.
<sup>3</sup>*n* = 13,085 individuals.
<sup>4</sup>High-risk variant carrier numbers in the Estonian Biobank subset (*n* = 17,679).

The 17 *BRCA1*/*2* variants comprised nine frameshift variants, six nonsense variants (including a novel predicted loss-of-function variant), and two missense variants. Thirteen (76%) variants were singletons. The most frequent KP variant was the known Eastern European founder mutation *BRCA1* c.4035delA (NM_007294.3),^34^ which accounted for 33% (*n* = 16/35) of *BRCA1* variant carriers. *BRCA1* c.5266dupC (NM_007294.3) was the second most prevalent KP variant,^34^ common throughout Eurasia,^35^ accounting for 23% (*n* = 11/35) of *BRCA1* variant carriers. The most frequently detected KP variant in *BRCA2* was c.8572C>T (NM_000059.3), seen in six (12.5%) carriers.

The novel, likely deleterious, *BRCA1* variant c.2178delT (NM_007294.3; p.Pro727Glnfs*9) causes a frameshift in exon 10 and creates a premature stop codon. The change is predicted to cause loss of normal protein function through nonsense-mediated decay of an mRNA due to the presence of a stop codon within the first ∼90% of the coding region. The variant is not present in ClinVar or dbSNP. As loss-of-function variants in *BRCA1* are generally pathogenic,^36^ we included c.2178delT (p.Pro727Glnfs*9) in the dataset as an LP variant. According to data from the Estonian Causes of Death Registry, the carrier of this variant was diagnosed with breast cancer under 50 years old and died of the disease.

### Participants with identified high-risk variants

Eight of the 48 participants with detected LP/KP *BRCA1/2* variants could not be re-contacted due to changes in residency status or death (*n* = 8). The cause of death for all deceased participants was cancer (Table S2).

Of the 40 contacted participants, 22 attended the first visit (55% response rate). One participant (a >75 year old woman) declined participation. For the 17 non-respondents (8 male, 9 female; mean age, 48 [range, 32–83] years), no information was available about the receipt of invitation letters or reason for non-response. One non-respondent had previously been diagnosed with breast cancer twice.

Of the 22 respondents (8 male, 14 female; mean age, 47.6 [range, 25–75] years), 18 had biological children (Table 2). Four (18%) respondents had previously received HBOC-related cancer diagnoses, and 18 (82%) respondents had first- or second-degree relatives with breast, ovarian, prostate, pancreatic, and/or endometrial cancer. One participant (ID.19) provided consent, but chose not to attend the second visit and did not receive genetic risk information.

**Table 2.** Characteristics of study participants and their relatives.

| ID | Sex | Age range | Gene with LP/KP variant | Personal ca.dx.and age range at dx | Family history<sup>1</sup> | 1st degree relatives with BC/OC/PC/PAC and age range at dx | 2nd degree relatives with BC/OC/PC/PAC and age range at dx | Cascades attended/suggested | BRCA1/2 variant positive cascades |
| --- | --- | --- | --- | --- | --- | --- | --- | --- | --- |
| 1 | F | 30-34 | BRCA1 | no | high | one relative (OC, 50-54) | no | 3 of 5 | 1 |
| 2 | F | 45-49 | BRCA2 | no | moderate | one relative (BC, 45-49) | one relative (PC, 75-79) | 2 of 5 | 2 |
| 3 | M | 65-69 | BRCA1 | PC, 55-59 | high | one relative (OC, 50-54) | four relatives | 0 of 5 | NA |
| 4 | F | 50-54 | BRCA1 | no | moderate | no | one relative (BC) | 2 of 5 | 1 |
| 5 | F | 35-39 | BRCA1 | no | high | one relative (OC, 55-59) | two relatives (BC, 55-59; OC, 40-44) | 4 of 7 | 1 |
| 6 | M | 40-44 | BRCA1 | no | moderate | one relative (BC, 50-54) | no | 0 of 3 | NA |
| 7 | F | 55-59 | BRCA1 | no | low | no | no | 5 of 8 | 2 |
| 8 | F | 75-79 | BRCA1 | BC, 75-79 | high | one relative (OC, 40-44) | no | 0 of 3 | NA |
| 9 | F | 75-79 | BRCA2 | BC, 50-54 | moderate | one relative (PAC, 55-59) | no | 3 of 8 | 0 |
| 10 | F | 30-34 | BRCA1 | no | high | one relative (BC, 35-39) | no | 1 of 7 | 1 |
| 11 | M | 75-79 | BRCA1 | PC, 60-64 | moderate | one relative (BC, 70-74) | no | 0 of 4 | NA |
| 12 | F | 45-49 | BRCA1 | no | moderate | no | one relative (BC, 55-59) | 1 of 2 | 1 |
| 13 | M | 40-44 | BRCA1 | no | high | two relatives (OC; PC 60-64) | no | 0 of 1 | NA |
| 14 | F | 30-34 | BRCA1 | no | moderate | no | tree relatives (BC, in their 50's) | 0 of 4 | NA |
| 15 | F | 40-44 | BRCA1 | no | moderate | no | one relative (BC, 50's) | 0 of 5 | NA |
| 16 | M | 25-29 | BRCA1 | no | low | no | no | 0 of 6 | NA |
| 17 | M | 55-59 | BRCA1 | no | moderate | no | one relative (BC, 50's) | 4 of 5 | 1 |
| 18 | F | 55-59 | BRCA1 | no | high | one relative (BC, 45-49) | no | 0 of 8 | NA |
| 19 | F | 25-29 | BRCA2 | no | high | two relatives (BC, 50-54; PC, 55-59) | two relatives (BC, 40's; PC, 60's) | NA<sup>2</sup> | NA |
| 20 | M | 40-44 | BRCA2 | no | low | no | no | 1 of 6 | 0 |
| 21 | M | 45-49 | BRCA2 | no | moderate | one relative (PC, 70-74) | no | 0 of 2 | NA |
| 22 | F | 30-34 | BRCA2 | no | limited | no | no | 0 of 1 | NA |

LP, likely pathogenic; KP, known pathogenic; BC, breast cancer; OC, ovarian cancer; PC, prostate cancer; PAC, pancreatic cancer.
<sup>1</sup>Per the National Comprehensive Cancer Network guidelines, high risk = one individual with two or more primary BCs, a first- or second-degree relative diagnosed with BC at $\leq 45$ years of age, two or more relatives with BC, male BC, or a relative with OC; moderate risk = some HBOC-related diagnoses, but insufficient for high risk qualification; low risk = no HBOC-related diagnosis reported.
<sup>2</sup>ID19 choose not to attend the second visit and did not receive genetic risk information, and was thus lost to follow-up.

Of the 102 first- and second-degree relatives for whom cascade screening was recommended, 26 relatives of 10 index participants (13 male, 13 female; mean age, 41 [range, 19–76] years) responded and attended (25.5% response rate; Table S3). No information is available about whether the non-responding relatives (55 female, 47 male) were informed or about their decisions to not participate. Nine cascade screening respondents were offspring, eight were siblings, and two were mothers of the index participants; the other seven individuals were second-degree relatives. Cascade screening identified 10 individuals as carriers of the family *BRCA1*/*2* variant. Four cascade-screened individuals had previously received different cancer diagnoses, being early-onset breast cancer in two cases. The response to cascade screening was higher through female than for male index participants (62% and 25%, respectively).

### Response to disclosure

All 21 participants who received results completed the first survey, and 13 of these participants completed the second survey (62% response rate; Table S4). The response rate was higher among women (85%) than among men (25%).

Twenty of 21 index participants reported that they were glad to have been contacted about the genetic findings; only one participant was unsure of how she felt about being contacted (Table 3, Q1). All participants considered the information provided to be understandable, interesting, informative, and valuable (Table 3, Q2–Q5). At 6 months, all 13 respondents reported that they were coping with the genetic information received and had no regret regarding their decision to receive it (Table 3, Q21–Q26).

**Table 3.** **Responses to surveys administered immediately and 6 months after disclosure of genetic risk information**

|  | Theme | Item | Statement | Mean | Range |
| --- | --- | --- | --- | --- | --- |
| Immediate response | **Decision regret<sup>1</sup>** | Q1 | I am glad that the Biobank contacted me regarding the genetic finding | 4.8 | 3-5 |
|  | **Perceived impact<sup>1</sup>** | Q2 | Information provided was understandable | 4.9<sup>2</sup> | 4-5 |
|  |  | Q3 | Information provided was interesting | 4.7 | 4-5 |
|  |  | Q4 | Information provided was informative | 4.9 | 4-5 |
|  |  | Q5 | Information provided was valuable | 4.9 | 4-5 |
|  |  | Q6 | I understand the potential impact of the finding on my close relatives | 4.9 | 4-5 |
|  |  | Q7 | I can explain the potential impact of this genetic finding on health risks to my family members | 4.7 | 4-5 |
|  |  | Q8 | I know who to turn to regarding health concerns or for counselling | 4.8 | 4-5 |
|  | **Emotional response<sup>3</sup>** | Q9 | I feel calm | 3.3 | 1-4 |
|  |  | Q10 | I am tense | 1.8<sup>2</sup> | 1-4 |
|  |  | Q11 | I feel upset | 1.8<sup>2</sup> | 1-4 |
|  |  | Q12 | I am relaxed | 3.5 | 1-4 |
|  |  | Q13 | I feel content | 3.3 | 1-4 |
|  |  | Q14 | I am worried | 2.2<sup>2</sup> | 1-4 |
| 6-month follow-up | **Emotional response<sup>3</sup>** | Q15 | I feel calm | 3.5 | 3-4 |
|  |  | Q16 | I am tense | 1.5<sup>4</sup> | 1-2 |
|  |  | Q17 | I feel upset | 1.7<sup>4</sup> | 1-2 |
|  |  | Q18 | I am relaxed | 3.3<sup>4</sup> | 2-4 |
|  |  | Q19 | I feel content | 2.9<sup>4</sup> | 1-4 |
|  |  | Q20 | I am worried | 1.8<sup>4</sup> | 1-3 |
|  | **Coping and decision regret<sup>1</sup>** | Q21 | I am able to cope with having this condition in my family | 4.8 | 4-5 |
|  |  | Q22 | It was the right decision | 4.8 | 4-5 |
|  |  | Q23 | I regret the choice that was made | 1.3<sup>4</sup> | 1-2 |
|  |  | Q24 | I would go for the same choice if I had to do it over again | 4.8<sup>4</sup> | 4-5 |
|  |  | Q25 | The choice did me a lot of harm | 1.3<sup>4</sup> | 1-2 |
|  |  | Q26 | The decision was a wise one | 4.7<sup>4</sup> | 4-5 |
|  | **Perceived impact<sup>1</sup>** | Q27 | I now have better access to health care / specialists | 3.5 | 1-5 |
|  |  | Q28 | I feel that my treatment and/or condition has improved | 2.8 | 1-4 |
|  |  | Q29 | The information received has somehow changed my life | 3.7 | 1-5 |

<sup>1</sup>Responses given on a five-point Likert scale: 5, agree; 4, slightly agree; 3, difficult to say; 2, slightly disagree; 1, disagree.
<sup>2</sup>Twenty of 21 participants responded.
<sup>3</sup>Responses given on a four-point scale: 4, very much; 3, moderately; 2, somewhat; 1, not at all.
<sup>4</sup>Twelve of 13 participants responded.

Immediately after receiving risk information with counseling, participants tended to feel content, calm, and relaxed (Table 3, Q9, Q12, and Q13), and only slightly or not at all upset, worried, or tense (Table 3, Q10, Q11 and Q14). However, the responses to these questions varied to a great extent. Overall, four individuals reported feeling tense and three reported feeling worried; one participant (ID.9) reported feeling tense, worried, and upset. Average response scores for these items were similar 6 months later (Table 3, Q15–Q20), although participants tended to feel less worried, upset, and tense than at the time of the first survey, with only one participant reporting feeling worried. Of the six individuals who reported feeling tense, worried, or upset immediately after receiving risk information, five responded to the 6-month survey and none of them reported having those negative feelings any longer. All 16 responding cascade screening participants reported that they were glad to have been contacted regarding the genetic finding in the family.

### Medical impact of result disclosure

Currently, the clinical management of study participants have been dependent on the personal or family history of HBOC-related cancers or national breast cancer population screening program (Table S5). One of the 22 families studied reported familial risk management by a clinical geneticist prior to the study. Based on their personal and family histories, eight (36.5%) of the 22 index participants qualified for HBOC genetic assessment, according to the NCCN criteria (Figure 2). The other 14 participants did not qualify for HBOC assessment according to the current guidelines because they had insufficient family histories or very limited information available. Four of 20 female carriers belonged to the age group covered by the national breast cancer screening program (Figure 2). Of the four women with histories of breast cancer, only one belonged to that age group at the time of diagnosis.

**Figure 2.**
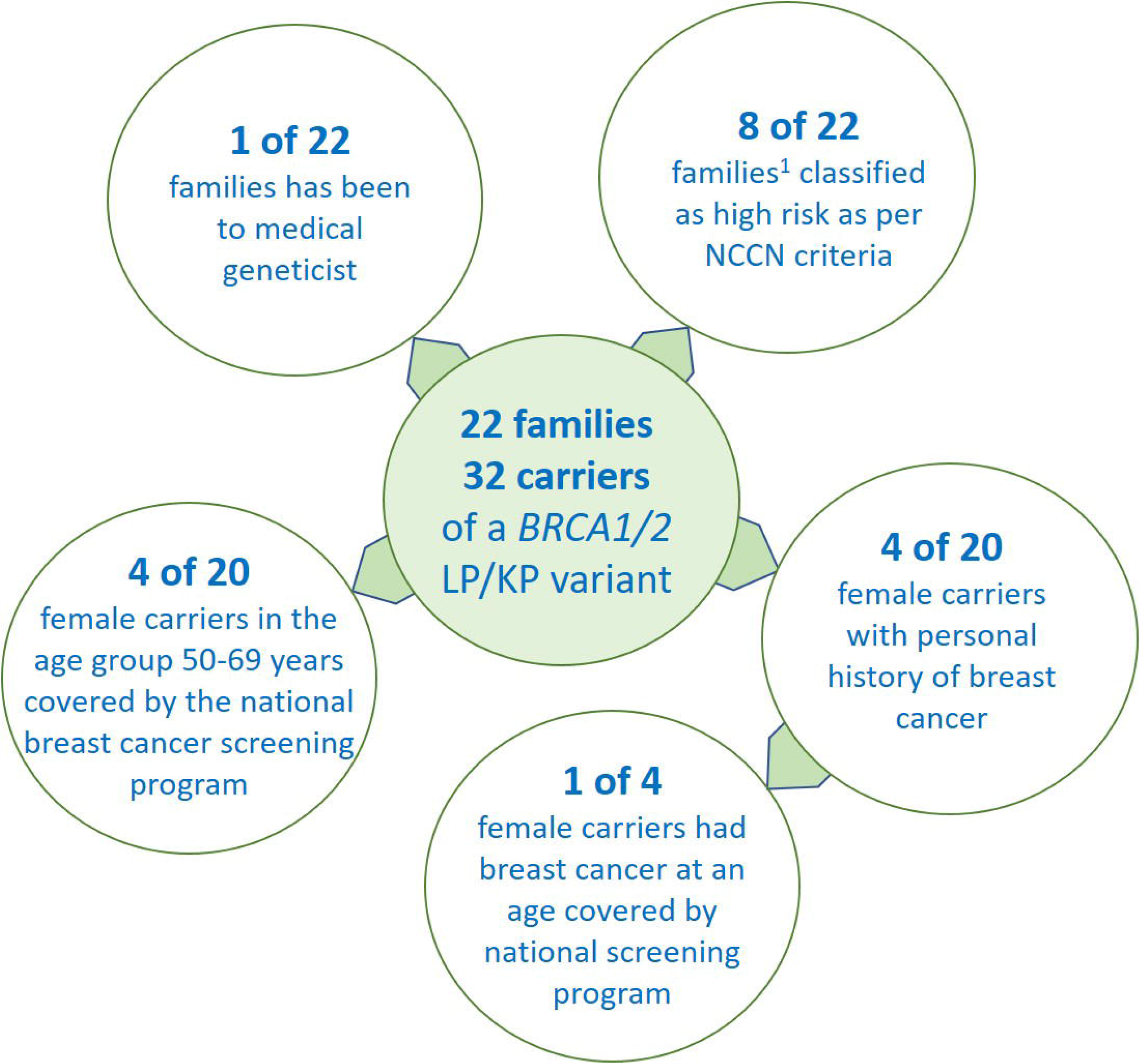
Added value of the genotype-first approach to the identification of high-risk individuals. The number of high risk individuals who have been to a medical geneticist is limited. Majority of the families do not meet the NCCN criteria to be classified as having high risk for HBOC. Only limited number of female carriers had breast cancer diagnosis at an age group covered by the national breast cancer screening program. ^1^Self-reported family history of hereditary breast and ovarian cancer–related cancers indicative of high risk.

All 19 women with *BRCA1/2* variants were referred to an oncologist for the development of personal surveillance plans. Two of these women were already seeing an oncologist due to previous cancer diagnoses. Over the 12–30-month follow-up period, 10 (59%) of these 19 women followed the clinical surveillance plan according to the HBOC guidelines (Table 4). Such adherence was not associated with participants’ age, whether they had children, family histories, or residence (rural or urban; proximity to a hospital).

**Table 4.**
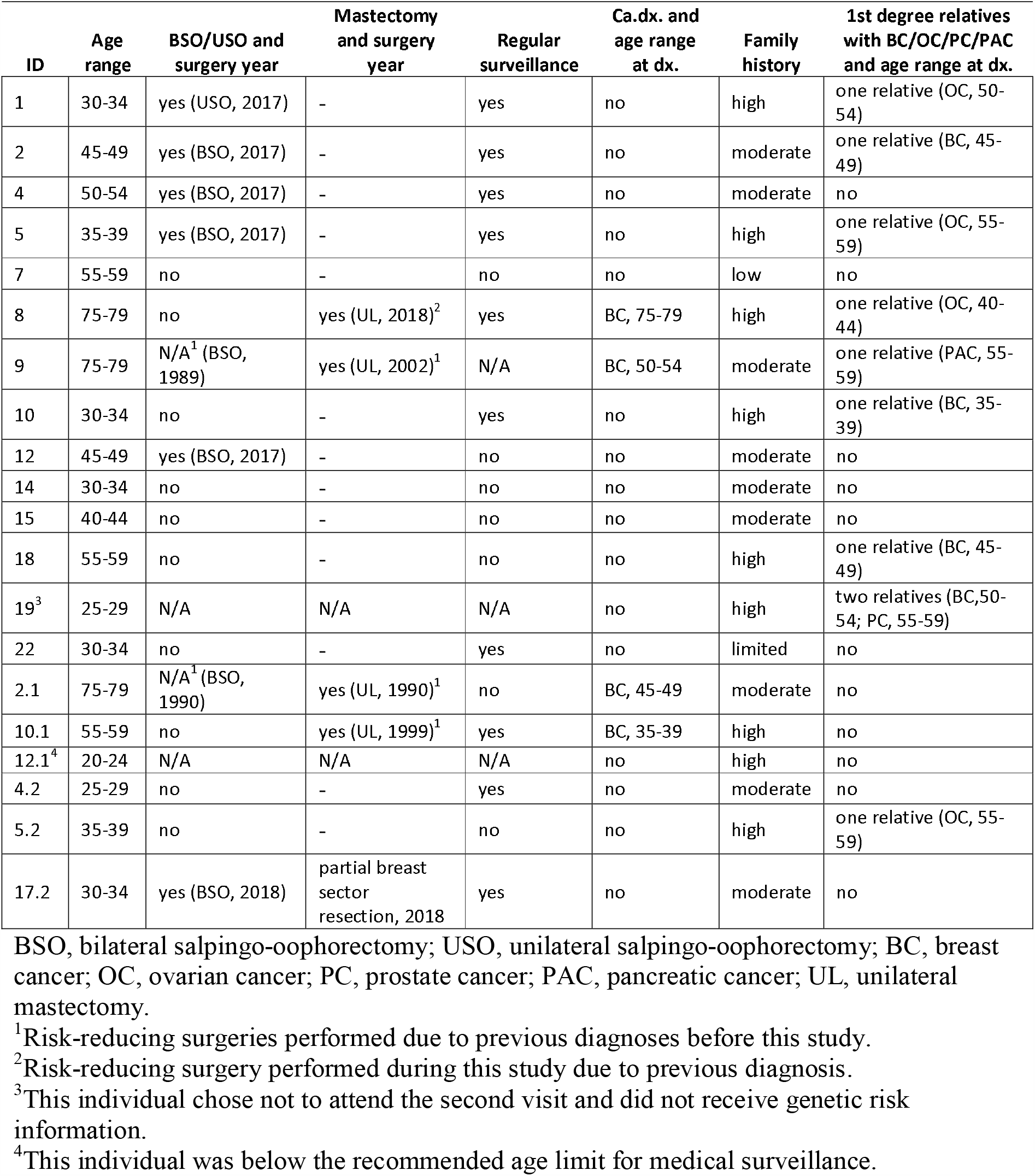
**Medical adherence among female index and cascade individuals with LP/KP *BRCA1*/*2* variants**

The NCCN guidelines for the management of individuals with high HBOC risk include recommendations for risk reduction surgery, depending on the subject’s age. Three of the 19 women with *BRCA1/2* variants had previously undergone unilateral mastectomy due to breast cancer. This includes two women with bilateral salpingo-oophorectomy (BSO) performed as part of empirical recurrence risk management. A <25 year old woman (ID12.1) was considered to be too young for regular clinical surveillance or preventive surgery. During the follow-up period, five (31.3%) of 15, eligible for preventive surgey, women chose to undergo preventive BSO with an average age of 43.6 (age range, 34–52) years. Additionally, from therapeutic interventions, one participant underwent unilateral salpingo-oophorectomy when a nonmalignant lesion was detected. Another participant underwent partial breast sector resection due to a finding after the study-related oncologist visit. No woman chose to undergo prophylactic mastectomy following the return of results. The eight women who did not choose to undergo risk-reducing surgeries during the follow-up period were aged 29–59 (mean, 40) years.

## DISCUSSION

This study highlights the significant potential of population-based genomic studies for personal risk evaluation and population-wide risk-based management at the national healthcare level. It provides critical evidence for the application of genotype-first screening to improve long-term outcomes for high-risk individuals in the population, many of whom are unaware of their genetic risk. Most importantly, they were not captured by current clinical practice, which emphasizes the importance of the population-based genomic screening.

### Potential of existing datasets

Population-based biobanks provide an excellent resource to study the frequency and penetrance of clinically significant genetic variants in unselected cohorts. A genotype-first approach to the analysis of unselected data yields an unbiased estimate of HBOC-related cancer prevalence in the general population, rather than solely in multiplex families. Intensive monitoring and early intervention can improve outcomes in carriers with no positive family history.^18^ The information on HBOC-related LP/KP variants affects the therapy of choice (e.g surgery approach, the use of PARP inhibitors in advanced cancer, etc.).^37^

The prevalence of LP/KP *BRCA1*/*2* variants in our study cohort was 0.80% (1/124), previous prevalence data for *BRCA1* (7.6%) and *BRCA2* (12.5%) was on clinical cohort^34^ of women with breast cancer diagnoses or predictive cases. The actual prevalence of *BRCA1*/*2* pathogenic variants is uncertain, as estimates are based largely on data from clinical cohorts, rather than general populations expected to be cancer free. The approach used in this study led to the identification of previously unknown carriers of LP/KP variants in *BRCA1*/*2*, irrespective of personal or family history.

Population biobanks are also a good resource for the identification of novel genetic variants that may be clinically significant. One novel, presumably pathogenic, *BRCA1* variant, c.2178delT (p.Pro727Glnfs*9), was identified in this study. The broad consent that biobank participants provide, the accessibility of biological sample collection, and the availability of long-term follow-up data in biobank datasets enable researchers to access and analyze information, including that from biobank participants who are deceased or cannot be contacted. The availability of multiple *in silico* pathogenicity evaluation scores and medical diagnoses enable estimation of the pathogenicity of novel variants, even when databases such as gnomAD, dbSNP, and ClinVar have no available data. Such variants may be population specific, yet still clinically relevant.

This study also revealed some limitations related to the approaching of high-risk individuals in the biobank setting. These limiting factors include difficulties with contact, as participants may have been contacted last more than a decade previously. Another challenge involved the composition of an invitation letter that respected participants’ right to not receive information while being sufficiently informative to allow participants to decide whether they were interested in participating in the present study. These factors could have contributed to the 55% response rate. The response rate to cascade screening (25.5%) was probably impacted, at least in part, by index individuals’ gateway roles. The response to cascade screening was higher for female than for male index participants, indicating the need for improved communication with gender-specific heritable disorders.^38,39^

The contacting of family members for cascade screening is one way to maximize the impact of a population biobank, and further investigation is needed to determine the best way to increase response rates for optimization of this cascade approach.

### Risk-based screening

As in many other countries, the national breast cancer screening program in Estonia targets women in a limited age group (50–69 years), thereby excluding some women who are potentially at high risk. In the present cohort, two women had received breast cancer diagnoses by the age of 50 years. Despite to the broad access to medical services in Estonia, the medical system consistently misses individuals with greater cancer risk. In our cohort, only one of 22 participating families had previously visited a clinical geneticist. These findings suggest that the majority of high-risk variant carriers are unaware of their genetic predisposition to HBOC-related cancers and are not under optimal surveillance.

The efficiency of breast cancer risk assessment based on family history depends on family size, family members’ ages, the sex distribution of high-risk genetic variants, and the availability of detailed individual knowledge of cancer in the family. For instance, our cohort contained three families with multiple diagnoses of cancer, but no information about primary disease locations. Additionally, the majority of family histories did not fulfill the current criteria for high risk.^17^

Genetic predisposition to cancer is usually suspected when the disease is diagnosed at a relatively young age. This pattern is largely true in familial cases seen in clinical settings, although the application of a genotype-first approach to an unselected population reveals great variability in the onset and prevalence of cancer. Our results indicate that age and family history alone are poor discriminators of breast cancer risk. We thus propose that existing genomic data be examined whenever possible during risk assessment.

### Risk-based management

The guidelines for women with suspected hereditary predisposition to HBOC recommend earlier and more frequent screening than regular age dependent screening and optional preventive surgery.^40^ In our cohort, 59% of female carriers followed the clinical surveillance recommendations during the follow-up period. Women with high-risk *BRCA1*/*2* variants tend to prefer surveillance for breast cancer. The uptake of risk-reducing surgeries in our cohort was lesser than reported previously, with 31% of our participants choosing to undergo BSO compared with the 46% reported by Rowley et al.^1^ and no participant choosing to undergo bilateral mastectomy. The uptake of risk-reducing procedures likely depends on different populations and cohorts, average age and other contextual factors including availability and reimbursement, clinical guidelines and traditions; these factors were currently not studied.

A possible limiting factor for long-term follow-up surveillance is that we obtained clinical follow-up data on individuals who participated in cascade screening from the collaborating oncologist and participating central hospital. Thus, these data may not reflect participants’ complete medical histories, including potential consultation with other oncologists and the use of surveillance services in the private sector. Thus, the actual rate of medical adherence may be somewhat higher than revealed by our analyses.

### Population-scale impacts of screening

Our analysis demonstrated that EstBB participants and their family members who participated in cascade screening appreciated receiving their screening results. This finding is likely generalizable to the general population.

Population biobank data provide information on the potential impact of population screening for high-risk individuals using tools currently offered to a limited group of individuals with significant personal or family histories indicative of genetic predisposition. The results of analogous genotype-first projects inform us about perceptions regarding the receipt of unexpected genetic results. Long-term follow-up data on high-risk variant carriers in the general population will aid assessment of the clinical utility of population screening.

At the end of 2019, more than 200,000 EstBB participants had been recruited. Array-based genotype data for all of these participants will become available by June 2020, and will be implemented as an integral part of the national personalized medicine initiative. Based on the 0.82% estimated prevalence of LP/KP variants from this study, more than 1,000 EstBB participants could be *BRCA1*/*2* variant carriers. Läll *et al*. suggests polygenic risk together with germline mutation testing could be an efficient complimentary tool for risk stratification in clinical practice for better screening and prevention.^41^ Considering a reduction in the detection rate due to the long-range phasing nature of array-based data and a response rate of ∼50%, the application of a genotype-first approach to HBOC and breast cancer screening in Estonia could impact a few thousand individuals, demonstrating the importance of this pilot evaluation of this type of approach.

## Data Availability

All results generated in the study are provided in the manuscript and as Supplementary Tables.

## ACKNOWLEDGEMENTS

We express our sincere thanks to the biobank participants and their relatives for participating in the study.

## CONFLICT OF INTEREST

Conflict of interest: none declared.

## FUNDING

This research was supported by the European Union through the European Regional Development Fund (project no. 2014-2020.4.01.15-0012 GENTRANSMED), European Union Horizon 2020 (grant no. 810645, no. 654248), Estonian Research Council (PUT736 to N.T., PUT PRG555 to N.T., IUT20-60, PUT1660 to T.E., PUTJD817 to M.K., MOBERA15, RITA1/01-42-03).

## Notes

### Competing Interest Statement

The authors have declared no competing interest.

### Author Declarations

The study protocol was approved by the Research Ethics Committee of the University of Tartu in 2016 (protocol no. 258/T-22 and no. 238/T-12)

